# Using Artificial Intelligence (AI) to Model Clinical Variant Reporting for Next Generation Sequencing (NGS) Oncology Assays

**DOI:** 10.1101/2025.05.14.25327648

**Authors:** Kenneth D Doig, Rashindrie Perera, Yamuna Kankanige, Andrew Fellowes, Jason Li, Richard Lupat, Ella R Thompson, Piers Blombery, Stephen B Fox

**Author notes:** **Corresponding Author:** Kenneth Doig. **EMAILS** Rashindrie Perera, Yamuna Kankanige, Andrew Fellowes, Jason Li, Richard Lupat, Ella Thompson, Piers Blombery, Stephen Fox.

## Abstract

**Background:** Targeted next generation sequencing (NGS) of somatic DNA is now routinely used for diagnostic and predictive reporting in the oncology clinic. The expert genomic analysis required for NGS assays remains a bottleneck to scaling the volume of patients being assessed. This study harnesses data from targeted clinical sequencing to build machine learning models that predict whether patient variants should be reported.

**Methods:** Three somatic assays were used to build machine learning prediction models (Stochastic Gradient Descent, Random Forest, XGBoost and Neural Networks). Using manual expert curation to select reportable variants as ground truth, we built models to classify clinically reportable variants. Assays were performed between 2020 and 2023 yielding 1,350,018 variants and used to report on 10,116 patients. All variants, together with 211 annotations and sequencing features, were used by the models to predict the likelihood of variants being reported.

**Results:** The tree-based ensemble models performed consistently well achieving between 0.891 and 0.995 on the precision recall/area under the curve (PRC AUC) metric when predicting whether a variant should be reported. To assist model explainability, individual model predictions were presented to users within a tertiary analysis platform as a waterfall plot showing individual feature contributions and their values for the variant. Over 30% of the model performance was due to features sourced from statistics derived in-house from the sequencing assay precluding easy generalization of the models to other assays or other laboratories.

**Conclusions:** Longitudinally acquired NGS assay data provide a strong basis for machine learning models for decision support to select variants for clinical oncology reports. The models provide a framework for consistent reporting practices and reducing inter-reviewer variability. To improve model transparency, individual variant predictions are able to be presented as part of reviewer workflows.

## INTRODUCTION

Clinical diagnostics in oncology has harnessed Next Generation Sequencing (NGS) to analyse patient DNA at the nucleotide level. This has allowed identification of diagnostic, prognostic and therapeutic information critical to a patient’s management. In earlier work^1-3^, we have highlighted limitations in scaling clinical sequencing, in particular, the effort required for the expert curation of the large number of somatic variants observed in a cancer patient’s DNA. Within a clinical laboratory, despite improvements in tertiary analysis software and databases, qualitative decisions about variants are frequently required by standard operating procedures (SOPs) and many subjective choices must be made while compiling a routine NGS clinical report. This often leads to inter-reviewer variability or the risk of clinically significant variants not being reported^4-7^. This study harnesses a corpus of over one million variants sequenced from 10,116 patients to build a number of AI models to support genomic scientists in identifying the variants that warrant curation. By using the expert curation and clinical reporting of 18,841 variants, we have built machine learning models that provide decision support tools for expert curators and provide objective, quantitative metrics to assess patient variants.

## METHODS

### NGS Reporting Workflow

This study used clinical patient data collected to provide diagnostic oncology reports. Samples for each assay were received and processed according to standard laboratory procedures and then sequenced. An automated bioinformatics pipeline was triggered at the completion of sequencing, which processed and annotated the sequencing data before loading the results into a tertiary analysis platform. Trained genomic scientists then analyse the variants and, based on quality control (QC) metrics and a large number of annotations, selected appropriate variants for reporting. Detailed descriptions of laboratory processes have been described previously^3^ and reviewed more recently^2^.

Sequenced variants were uploaded to either an in-house tertiary analysis decision support software system called PathOS^3^ (‘Haem’ and ‘Myeloid’ assays) or a commercial analysis platform, PierianDX CGW (Solid Tumour), for filtering, analysis and reporting. Reported variants were manually curated to establish variant action within a patient’s clinical context using either ACMG or AMP guidelines or other custom classifications^8,9^. Curated variants with enriched expert annotations were deposited within a common database enabling subsequent patients presenting with the same variants to be matched to the existing variant annotations so that prospectively; only novel variants need be curated. The patient’s clinical context was also stored with curated variants to inform decisions on whether the same variant appearing in a different clinical context warrant using the same stored curation or whether a new distinct, and perhaps adapted, curation of the variant and context would be required. For details of the pipelines and curation workflows please refer to previously documented processes^1-3^ and the Supplementary Methods section.

### Data Extraction

This study used clinical patient data collected over a period of four years within the Pathology Department at the Peter MacCallum Cancer Centre (see Table 1).

**Table 1.** NGS Assays Characteristics Used In Study.

| Abbreviation | Assay | Runs | Patients | Total Variants | Unique Variants | Reported Variants | Unique Reportable | Reportable Genes |
| --- | --- | --- | --- | --- | --- | --- | --- | --- |
| TSO500 | Illumina TruSight Oncology 500 | 195 | 1,772 | 804,074 | 33,809 | 5,112 | 2,514 | 249 |
| Haem | QIAGEN QIAseq custom panel | 157 | 5,815 | 444,073 | 20,126 | 11,816 | 4,554 | 57 |
| Myeloid | QIAGEN QIAseq custom panel (filtered gene list) | 157 | 2,529 | 101,871 | 4,079 | 1,913 | 774 | 8<br>(Oct21-Jun22)<br>or 22<br>(Jul22-Dec23) |
|  |  |  | 10,116 | 1,350,018 |  | 18,841 |  |  |

Variant data was extracted from tertiary analysis systems (PathOS or PierianDX CGW), and a number of other annotation sources were used to enrich the data using data source APIs (MyVariant, Genome Nexus) or downloaded datasets (Cosmic Cancer Mutation Census^10^, DeepMind AlphaMissense^11^, MSK Cancer Hotspots^12,13^). The annotation sources were matched to the tertiary analysis data using the GRCh37 genomic representation of the variant (chromosome, position, reference bases, alternate bases). Both the MyVariant and Genome Nexus sources aggregate multiple third -party data sources providing annotation efficiency. MyVariant provided access to the sources dbNSFP^14^, CADD^15^, CIViC^16^, ClinVar^17^ and gnomAD^18^ while Genome Nexus was used to access OncoKB^19^, SignalDB^20^ and dbSNP. To match downloaded Cosmic Mutation Census data, the sequenced dataset was queried by the HGNC gene and HGVS coding (HGVSc) format of the variant nomenclature.

The resulting annotated variant set used for the machine learning ingestion process comprised 1,350,018 rows and 211 columns. The split between the three project assays is shown in Table 1. The refined subset of annotations (features) used for the models is described in Table 2.

**Table 2.**
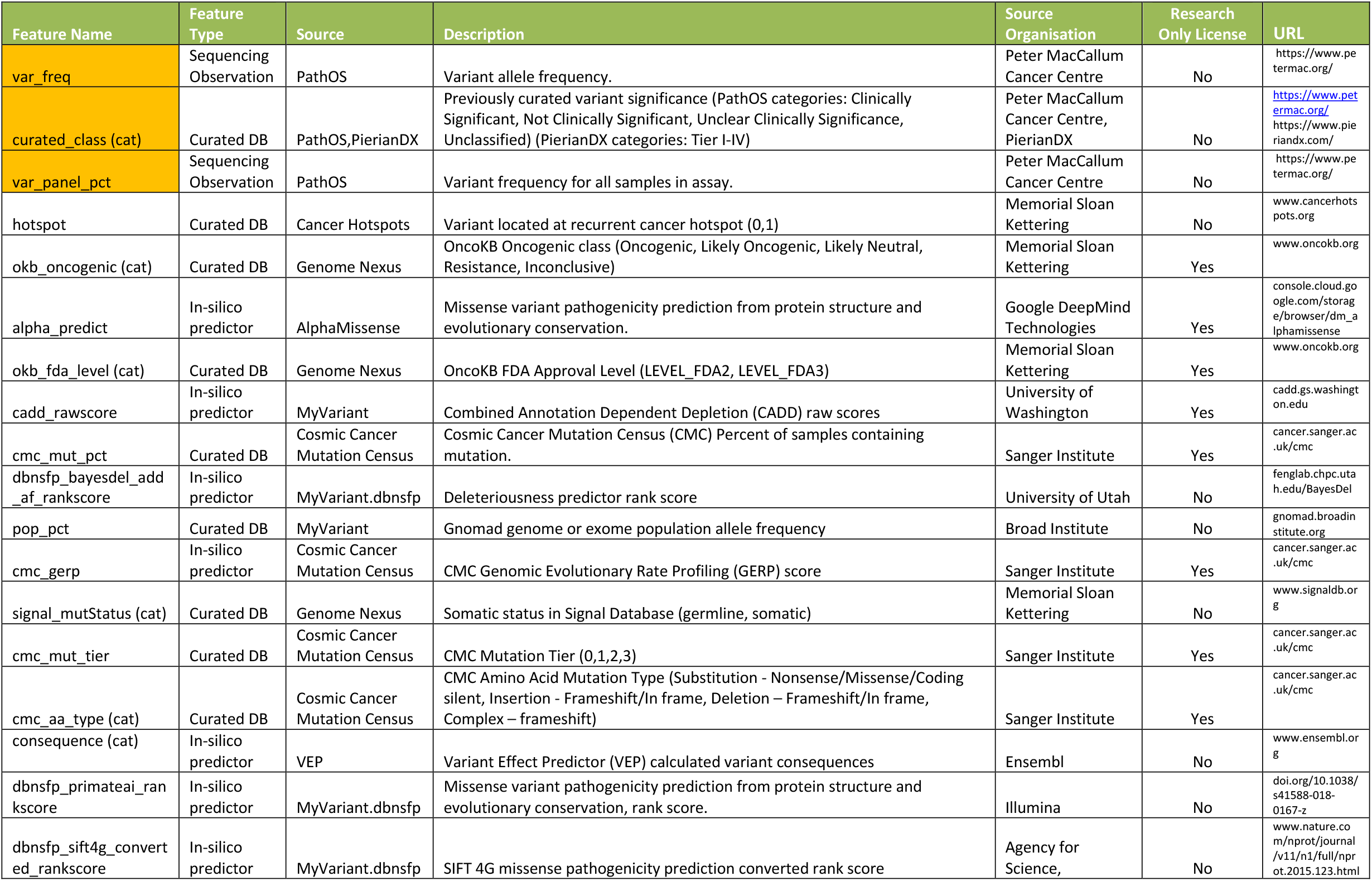
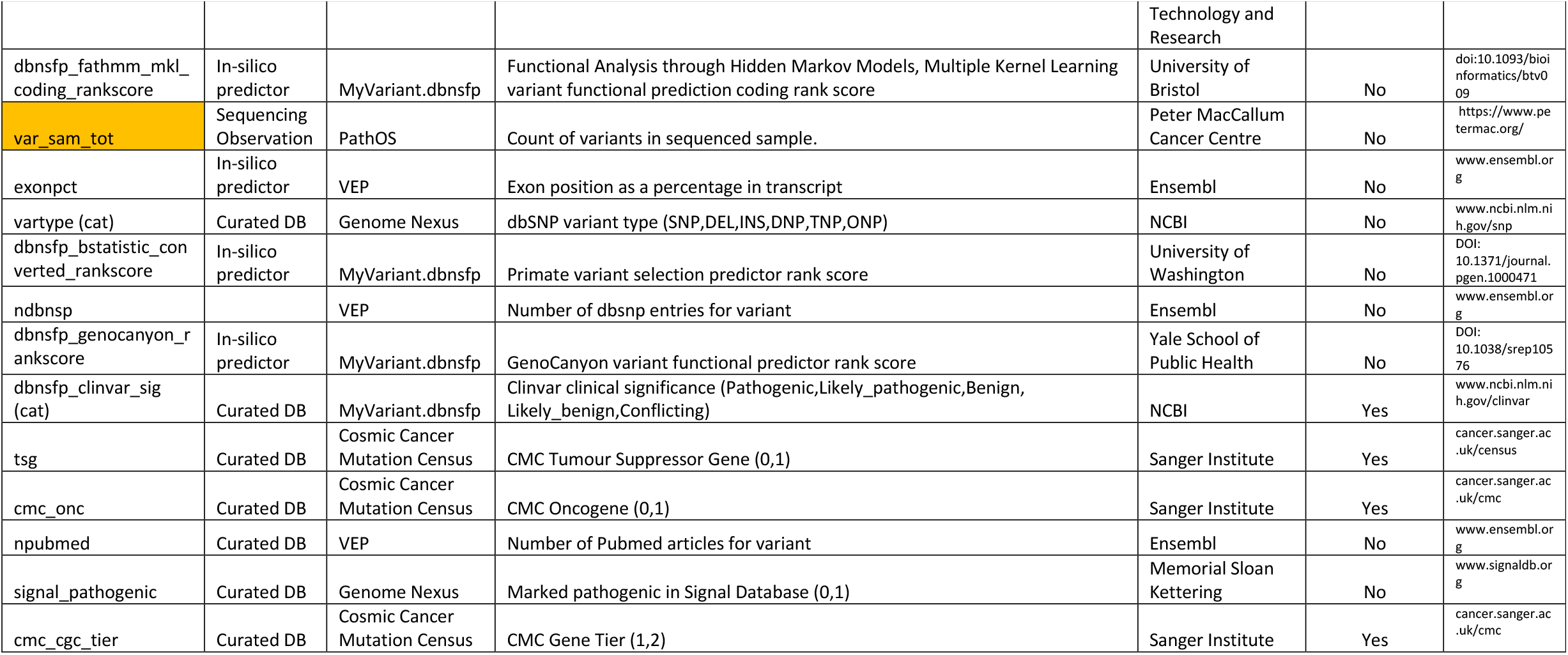
Feature set used for final models. In-house features highlighted in orange. All features are numerical except for categorical features marked “(cat)”.

To emulate the steps used by many laboratories to enrich variants for relevant somatic mutations, the following filters were applied to the full dataset.

1. Removal of variants with predicted consequences (synonymous, intronic, 5’ UTR) unless they also were predicted to be a splice variant (remove variants without protein coding consequences),
2. Removal of variants with a variant frequency less than 2% (remove variants occurring towards the assay limits of detection),
3. Removal of variants occurring at greater that 1% in the population (frequently occurring variants are unlikely to be deleterious),
4. Removal of variants occurring in more than 35% of the samples within the assay (frequently occurring variants are likely to be assay artifacts or frequently occurring in the population).

Both the full set of variants and the filtered variant dataset were used to build and compare machine learning models.

To compare the relative importance of features by assay, the Scikit-learn Random Forest feature importance property was used which provides an impurity-based score per feature summing to 1. For categorical features, individual category values within a feature were summed to give a single feature value.

## METHODS (CONT.)

### Machine Learning

#### Data Ingestion and Pre-processing

Data processing was employed to ensure suitability for model fitting. The raw data was filtered to identify numerical and categorical features. Irrelevant feature columns, including HGVS formatted variants, were removed from the categorical data set. Two separate preprocessor pipelines were then employed to handle the two different data types. For numerical data, missing values were imputed with a constant value of 0. Scikit-learn’s RobustScaler was then used to normalise these features. RobustScaler removes the median and scales the data according to the quantile range, effectively reducing the influence of outliers on the model’s learning process. Missing values in categorical features were imputed with a dummy value not occurring within that feature. It was not appropriate to drop rows with missing values as the annotation databases used to enrich the variant features often had limited coverage of the observed sequence variants. The imputed categorical data was then converted into numerical representations using Scikit-learn’s OneHotEncoder. This encoder replaces categorical features with a set of binary features, one for each distinct category value.

#### Model Estimators

To explore a range of learning algorithms and identify the most effective models for predicting variant reporting, four diverse machine learning models were employed:

- Stochastic Gradient Descent Classifier (SGD): This model fits a linear Support Vector Machine (SVM) using a Stochastic Gradient Descent optimiser. SGDClassifiers are known to be able to handle large datasets efficiently.
- Random Forest (RF): This tree-based ensemble method combines multiple decision trees, where each tree makes a prediction, and the final prediction is the majority vote from all trees. RF is known for its robustness to noise and ability to handle complex relationships within the data.
- Extreme Gradient Boosting (XGB): This powerful tree-based ensemble method leverages gradient boosting to iteratively learn from the training data, focusing on areas where previous models made errors. XGBoost is known for its accuracy and ability to handle complex features.
- Neural Network (NN): This flexible model learns complex, non-linear relationships between features and the target variable. A multi-layer perceptron architecture with backpropagation was used for training. NNs can be particularly effective in capturing intricate patterns within the data.

By utilizing this diverse set of models, a broad range of learning styles was captured to identify the approach that generalises best to unseen data for accurate variant clinical reporting prediction.

#### Model Estimator Hyperparameters and Architectures

The parameters and the architecture of the models were chosen empirically. The SGD classifier employed default hyperparameters, while the Random Forest classifier was configured with 300 decision trees (n_estimators=300) to create a robust ensemble model. The XGBoost regressor was configured with 1,000 estimators (n_estimators=1000) and a maximum depth of 20 (max_depth=20) to enhance its ability to capture complex relationships within the data. All three models utilised a constant random state to ensure consistent results across training runs.

A multi-layer perceptron architecture was implemented for the NN model. The architecture utilises a sequential stack of densely connected layers with ReLU (Rectified Linear Unit) activation functions for non-linearity. The first hidden layer has 50 neurons, followed by two additional hidden layers with 6 neurons each. Dropout layers with a rate of 0.4 are incorporated after the first two hidden layers to minimise overfitting. The final output layer has a single neuron with a sigmoid activation function, generating a probability value between 0 and 1 representing the predicted likelihood of a variant requiring clinical reporting. The Adam optimiser was used for efficient training, and the binary cross-entropy loss function was employed to measure prediction error during the training process. Additionally, early stopping with a patience of 1 was employed to stop training once the validation accuracy reaches the maximum to avoid overfitting on the training data.

#### Feature Pruning

To identify the most informative features for variant clinical reporting prediction, recursive feature elimination (RFE) was employed. RFE iteratively removes the least important feature based on its ranking in the previous iteration. This process continues until the desired number of features (40 in this case) is reached. For feature selection, we employed a RF model with n_estimators set to 50. This number was chosen based on preliminary evaluations, which demonstrated that 50 estimators effectively allowed us to obtain stable feature rankings without incurring excessive computational costs. By focusing on the most relevant features, RFE helps improve model performance and reduces the risk of overfitting by preventing the model from relying on irrelevant information.

#### Model Evaluation

To create training and testing sets for evaluating model performance, we created a custom variant of the Scikit learn TimeSeriesSplit cross validator^21^. Batched sequencing run analysis is temporal in nature and uses data from the expert curated variants of earlier runs. It is critical to only validate the models using sets of variants from sequencing runs occurring after the runs used in the training set. K cross validation folds were created by partitioning the variants by splitting runs into K+1 sets. The Nth cross validation fold uses the earliest N partitions as the training set and the N+1 partition as the test set. Overall, all data is used for both training and testing, preventing data leakage, and ensuring a realistic model evaluation. This approach recapitulates the time-based process where genomic analysis reuses the analysis of previous sequencing run analysis.

Model performance is often measured with F1 scores or Receiver Operating Characteristics (ROC) plots, but for strongly imbalanced dataset as frequently found in health studies ROC plots are less useful. Here the number of positive results, (reportable variants), greatly outnumbered the negative results (benign or technical artefact variants), so precision recall curves (PRC) and their associated area under the curve (AUC) were used for model performance metrics^22^.

## RESULTS

The tree ensemble estimator models (see Table 3), performed consistently well across the assays achieving between 0.891 and 0.995 for XGBoost and between 0.907 and 0.995 for Random Forest on the precision recall area under the curve (PRC AUC) metric. Both the linear SGD model and the Neural Network model performed less consistently and poorly on the Myeloid assay with its smaller datasets. The test sets were chosen as described in the Model Evaluation section.

**Table 3.**
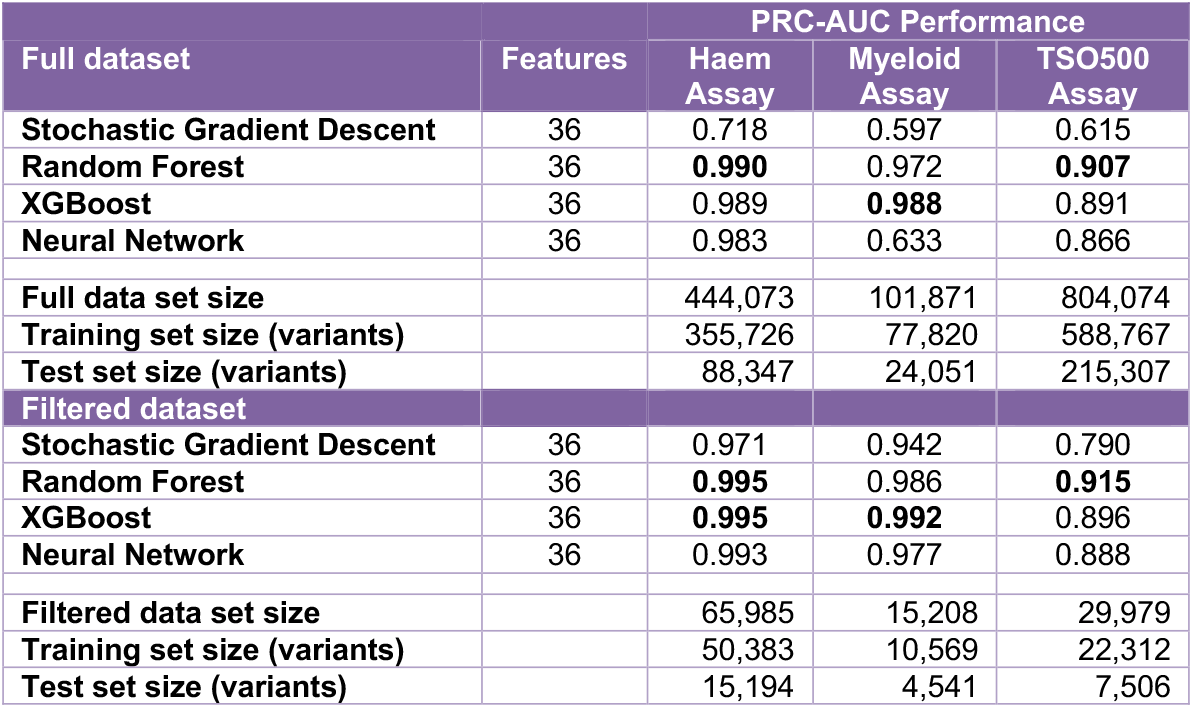
Model Performance on approximately 20% hold out test set.

| Full dataset | Features | PRC-AUC Performance |  |  |
| --- | --- | --- | --- | --- |
|  |  | Haem Assay | Myeloid Assay | TSO500 Assay |
| Stochastic Gradient Descent | 36 | 0.718 | 0.597 | 0.615 |
| Random Forest | 36 | **0.990** | 0.972 | **0.907** |
| XGBoost | 36 | 0.989 | **0.988** | 0.891 |
| Neural Network | 36 | 0.983 | 0.633 | 0.866 |
| Full data set size |  | 444,073 | 101,871 | 804,074 |
| Training set size (variants) |  | 355,726 | 77,820 | 588,767 |
| Test set size (variants) |  | 88,347 | 24,051 | 215,307 |
| Filtered dataset |  |  |  |  |
| Stochastic Gradient Descent | 36 | 0.971 | 0.942 | 0.790 |
| Random Forest | 36 | **0.995** | 0.986 | **0.915** |
| XGBoost | 36 | **0.995** | **0.992** | 0.896 |
| Neural Network | 36 | 0.993 | 0.977 | 0.888 |
| Filtered data set size |  | 65,985 | 15,208 | 29,979 |
| Training set size (variants) |  | 50,383 | 10,569 | 22,312 |
| Test set size (variants) |  | 15,194 | 4,541 | 7,506 |

The representative profile of the ROC and PRC data are shown in Figure 1.

**Figure 1.**
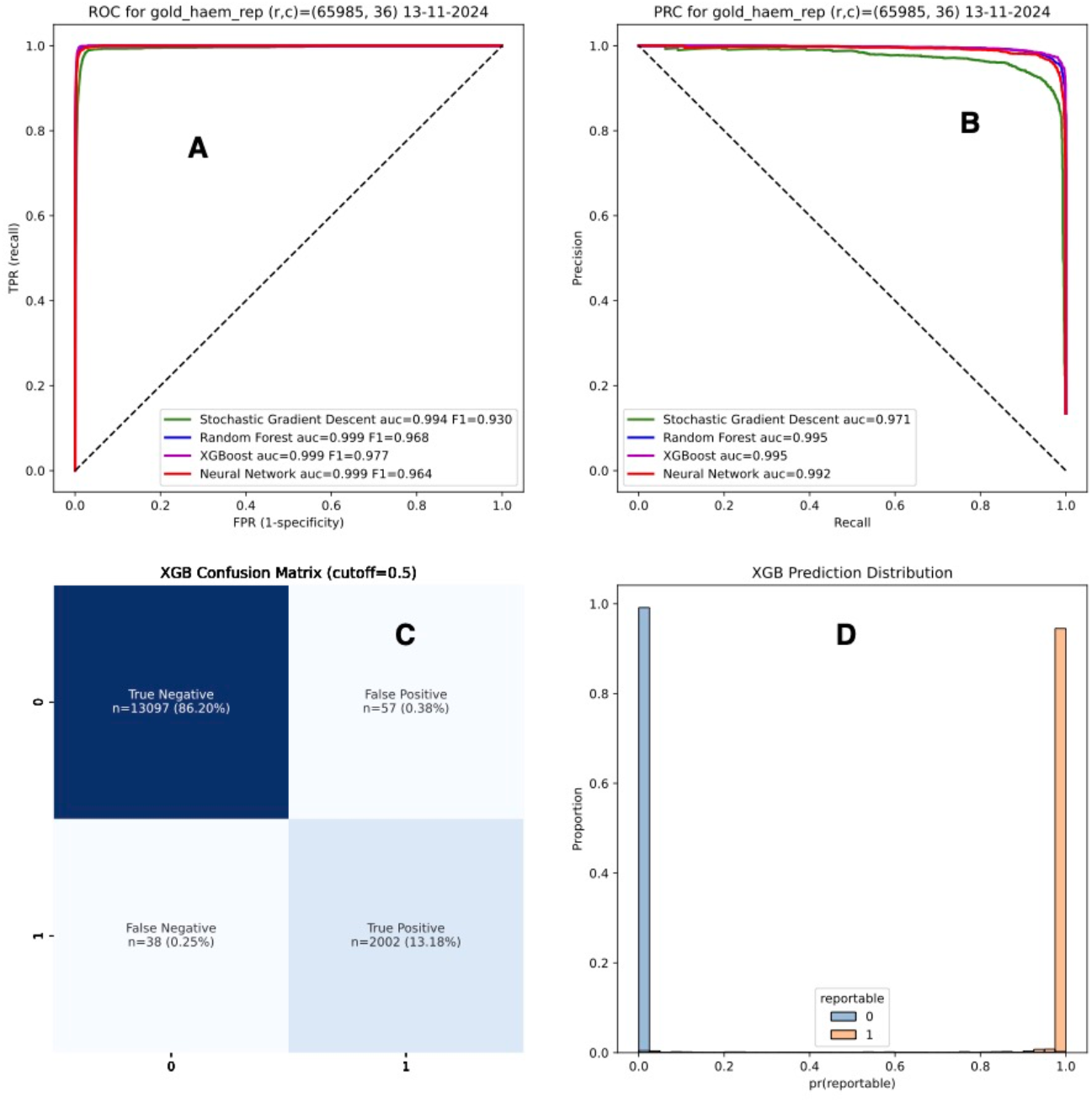
Model estimators performance for the filtered Haem dataset. (A) Receiver Operator Curves (ROC) and (B) Precision Recall Curves (PRC) for the Haem assay and all model estimators. Performance is on a hold out test set (n=15,194) and model training set (n=50,383). (C) Confusion matrix with a 0.5 cutoff for the XGBoost estimator. (D) Binned prediction probabilities for the XGBoost estimator.

Time series cross validation was performed as described in section Model Evaluation. Figure 2 shows the performance for the XGBoost estimator. As the training set size increases with each CV fold, the PRC AUC can be seen to plateau, particularly for the TSO500 solid tumour assay, whose performance is below that of the haematological assays. This performance difference is likely due to the mean number of reportable raw variants relative to reportable variants per patient: TSO500 (total=453.8, reportable=2.9 (0.6%)), versus the Haem (total=76.4, reportable=2.0 (2.6%)) and Myeloid (total=40.3, reportable=0.8 (2.0%)) assays.

**Figure 2.**
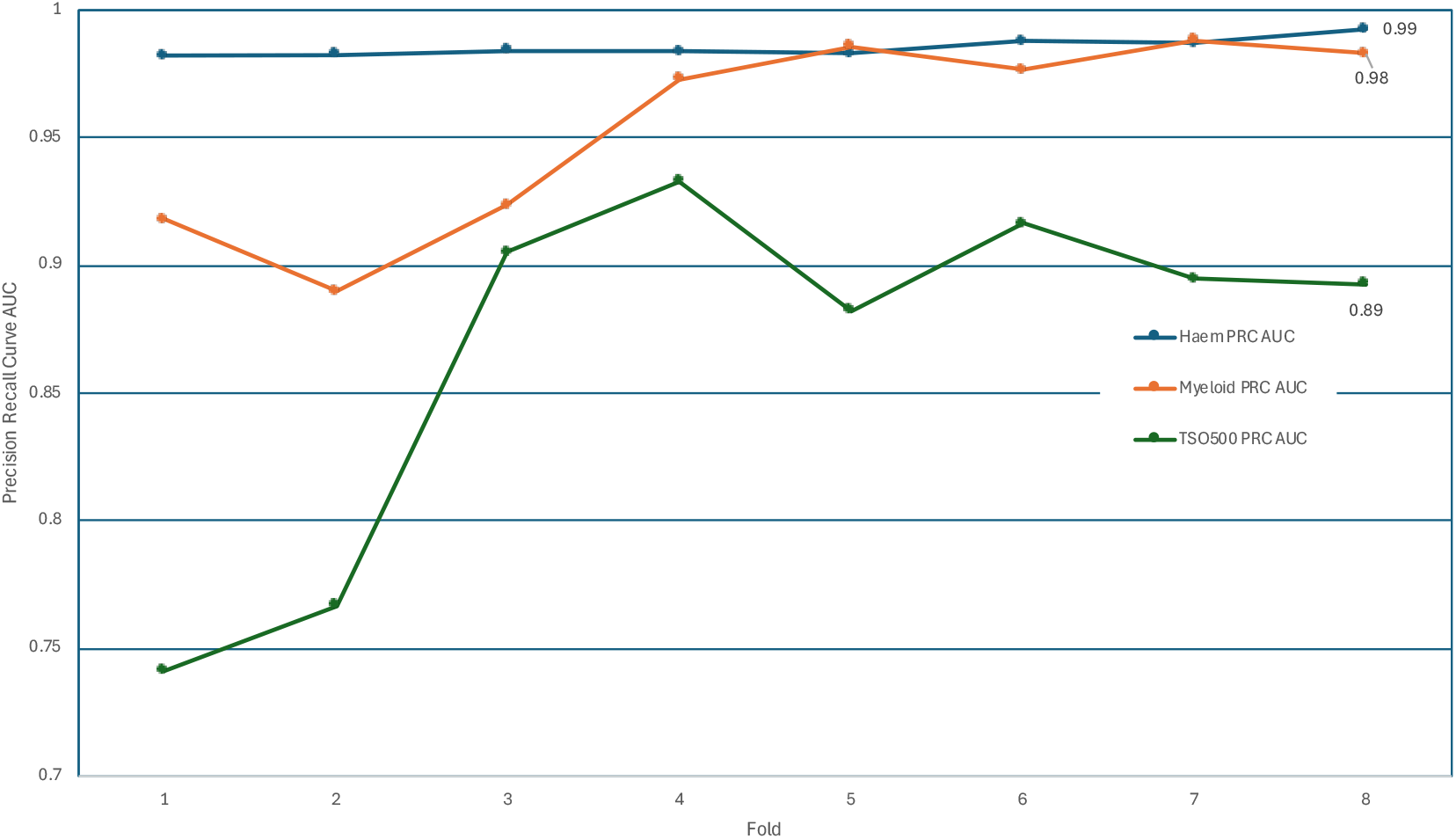
Assay time series 8-fold cross validation performance. Precision Recall Curves AUC values for each assay for the XGBoost estimator.

In addition, the percentage of novel variants for the TSO500 assay is approximately double the percentage for that of the Haem and Myeloid assays (see Figure 3). The number of novel variants progressively decreased as the in-house knowledgebase accumulated data on more reportable variants over time. Up until June 2022, the Myeloid panel had 8 reportable genes, this increased to 22 reportable genes in July 2022 leading to the early variability of the Myeloid data points in Figure 3. Both the Haem and the Myeloid assay graphs start in October 2021 but benefit from reported variants curated prior to this date and therefore have a lower number of novel variants initially.

**Figure 3.**
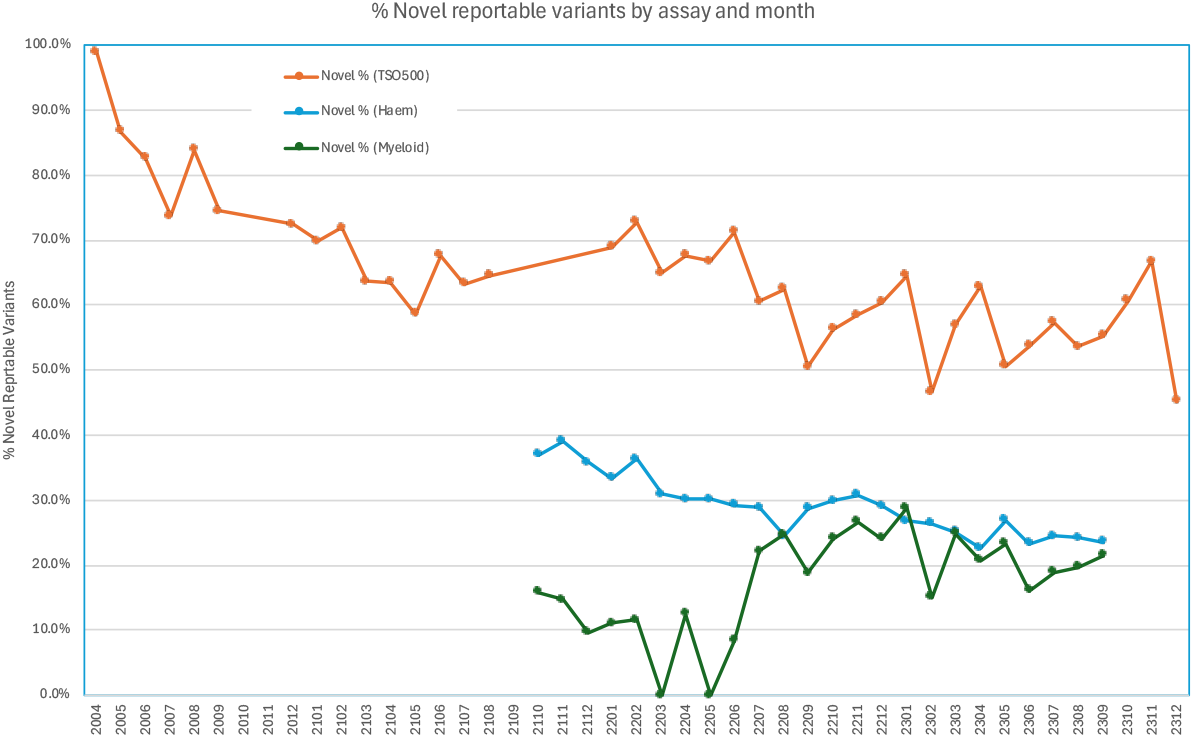
Percentage of Novel Variants by Assay and by Month. The importance of features used in the models were calculated as discussed in the Data Extraction section. The relative importance for the Haem assay is shown in Figure 4. and the feature names, their source and description are shown in Table 2. The features have been placed into four groups as follows; Sequencing and assay specific features (Seq), curated knowledge bases, both public and in-house (KB), reference genome variant effect (G) and public *in-silico* predictors (IS).

**Figure 4.**
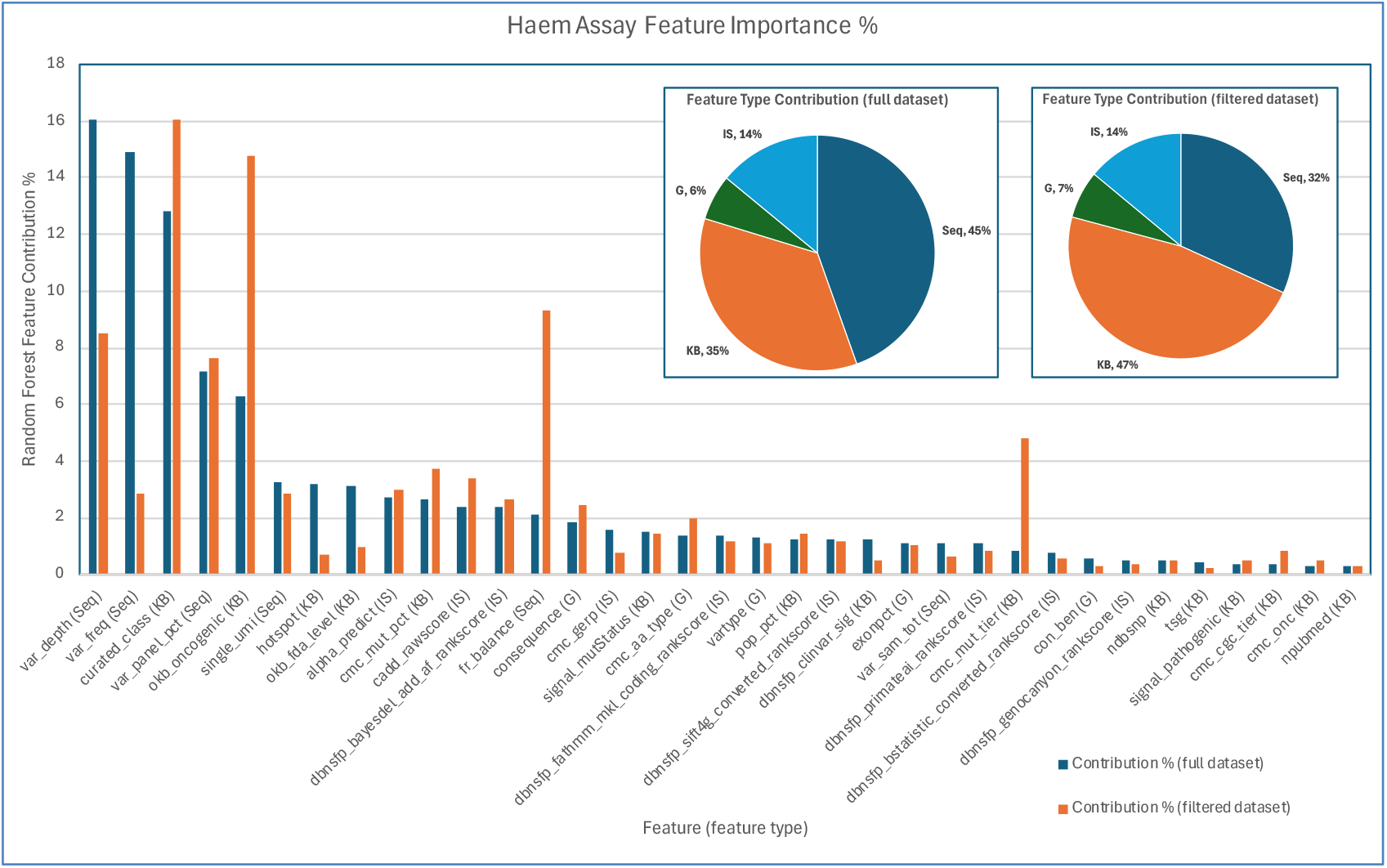
Haem Assay Feature Importance. Feature importance (%) for the Random Forest model for the full Haem Assay dataset (green bars) and filtered dataset (orange bars). Features are annotated by feature type in parenthesis; sequencing and assay specific features (Seq), curated knowledge bases, both public and in-house (KB), reference genome variant effect (G) and public *in-silico* predictors (IS). The inset pie charts show the relative contribution of feature types for both the full and filtered datasets.

Model inference of variants from new sequencing runs can provide decision support for genetic scientists interpreting patient samples. For our clinical model implementation of individual variant predictions, we selected non-linear tree-based models that provide a quantitative interpretation of the contribution each model feature adds to the final prediction for each variant. Tree-based models such as Random Forests^23^ and XGBoost^24^ lend themselves to detailed interpretations of predictions using Shapley values, an approach from cooperative game theory^25^. The interpretation of the additive probability contributions of features can be most naturally represented as a waterfall plot where the features are sorted from top to bottom in order of decreasing contribution effect. See Figure 5.

**Figure 5.**
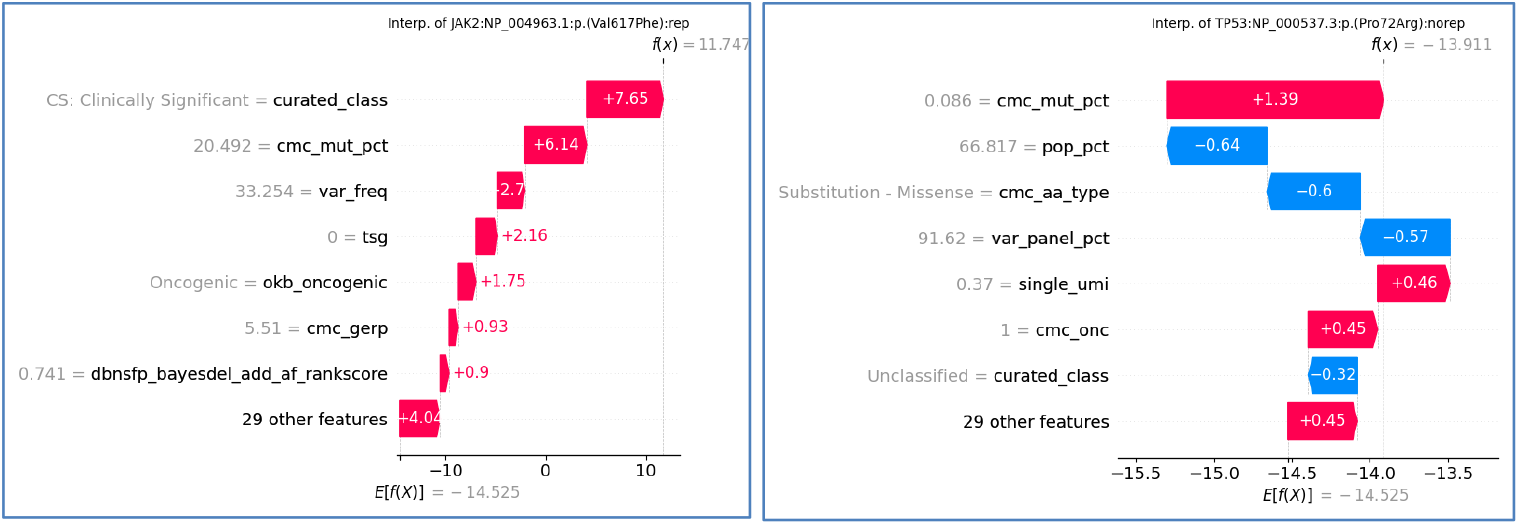
Variant interpretation waterfall plots. (a) Interpretation plot for variant JAK2 (Val617Phe) predicting it should be clinically reported by the model with high scoring features of curated class and Cosmic Cancer Mutation Census. Features contributing to this prediction are shown in order of weight of contribution from largest (top) downwards. The annotation values of each feature are shown in grey to the left of the feature name. Red indicates a feature that positively contributes to a variant being reported while blue indicates a negative contribution. (b) Waterfall plot for the variant TP53 (Pro72Arg) showing its high prevalence in the general population (pop_pct=66.8%) and its frequent occurrence in the panel samples (var_panel_pct=91.6%) give a low prediction of it being reportable.

The interpretation plots are made available to the genomic scientist performing variant analysis through the in-house tertiary analysis platform (PathOS^3^). This allows patient variants to be sorted by model prediction likelihood and the viewing of individual variant model interpretation through a popup window displaying how the model used variants attributes. See Figure 6.

**Figure 6.**
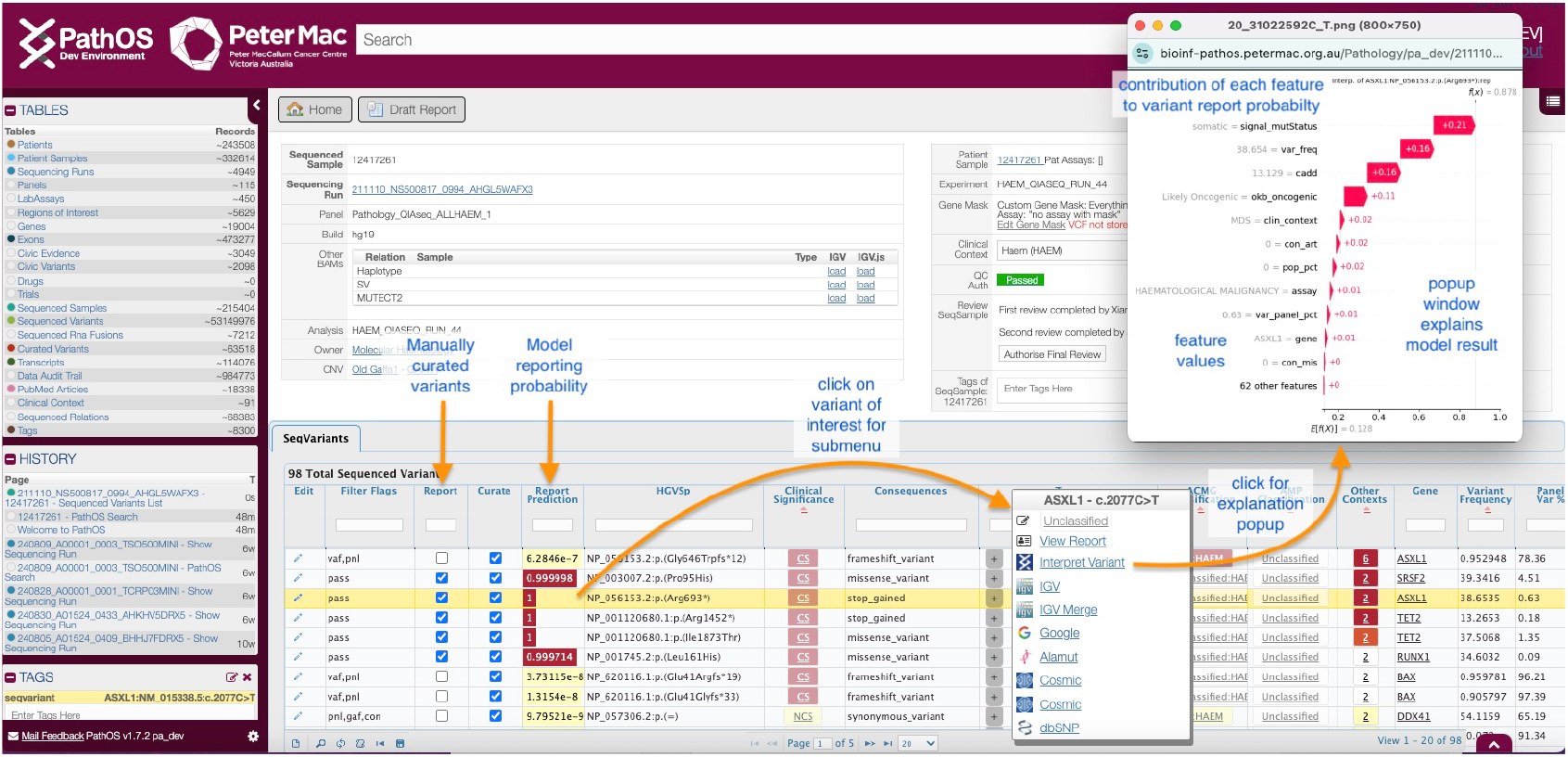
Model Integration with Tertiary Analysis Platform (PathOS)

## DISCUSSION

This study created machine learning models to assess suitability for providing decision support for clinical reporting of NGS targeted assays. The performance of tree-based ensemble models (Random Forest and XGBoost) on all assays ranged from 0.907 up to 0.995 PRC AUC. There was an incremental improvement (full dataset mean=0.962, filtered dataset mean=0.9667) when using the filtered data that used cutoff values to remove variants unlikely to be diagnostically relevant for reporting. Filtering variants greatly reduced the volume of unreported variants while maintaining reportable variants and allowing the models to focus on identifying reportable variants rather than excluding unreported variants. While providing a slight improvement in performance, filtering with hard cutoff criteria risks excluding variants that may be reportable near the limit of cutoff values. For both the full and filtered datasets, the solid tumour (Illumina TSO500) assay did not perform as well as the haematological assays (Figure 2,Table 3) although the model performances (PRC AUC of full dataset 0.907 and filtered dataset 0.915) are still likely to provide a significant benefit for genome analysts.

Comparison of which features drove model performance showed clear differences between models built on the full datasets compared to the filtered variant data. Features derived from the sequencing process (variant depth, variant frequency *etc*.), are the most important feature type (Seq=45%) compared with knowledge-based features (KB=35%) for the full Haem dataset. In contrast, for the filtered dataset, the situation is reversed (Seq=32%, KB=47%). This can be understood by noting the sequencing-based features are used to filter out technical artefacts from the NGS process that are dominant in the full datasets. In the filtered dataset, where many of these variants are removed, the model more fully utilises features from variant knowledgebases to identify reportable variants. For the Haem assay filtered dataset, the knowledge base features of most importance were: curated_class 16.1% (in-house curation DB), okb_oncogenic 14.8% MSK OncoKB and cmc_mut_tier 4.8% Cosmic Cancer Mutation Census. For the TSO500 assay filtered dataset, the two top features where okb_oncogenic 15.4% and signal_mutstatus 9.0% MSK Signal DB. It is noteworthy that of these data sources, Memorial Sloan Kettering’s Signal database is the only one available for non-research use. The usage restrictions on large curated variant databases are an emerging issue for clinical laboratories wishing to improve their curation processes. The large contribution of in-house assay sequencing features (Seq) to model performance (full dataset: 45%, filtered dataset: 32%) represent features that are difficult to be generalised to other assays. In addition, these features may not generalise easily between laboratories running the same assay due to differences in SOPs, sample preparation and local conditions.

The dependency on sequencing-based features for all models built from the full dataset or filtered datasets suggests that generalising these types of models between laboratories or assays will be difficult. The models depend on features and statistics derived from the actual sequencing process (feature group Seq) which are not present public variant databases (see Figure 4).

A key weakness of many initiatives to introduce AI into healthcare stem from the adoption of opaque or ‘black box’ models which don’t make transparent the interpretation of individual predictions. This issue has been raised multiple times in the literature^26,27^, and it is becoming less acceptable for models in healthcare to only predict outcomes without explanation. Aggregated model feature importance is commonly available but local interpretation of individual predictions is more difficult. Understanding individual predictions is necessary for trust, accountability and to confidently action model results. Individual prediction interpretations have been classed as a *mandatory criteria* in the recently released AI in Healthcare Guidelines released by the Dutch Ministry of Public Health^28,29^. This has also been driven by the European GDPR and other regulations which further encourage a ‘right to explanation’ for AI systems^30^. Individual variation interpretations can also reduce Automation Bias (AB), the tendency of over relying on automation^31^.

## CONCLUSION

The implementation of machine learning models in the clinical reporting of NGS assays is emerging as AI is applied to variant interpretation in oncology. This study has shown data derived from targeted NGS assays can serve as a robust base for generating predictive models that support genetic scientists in clinical decision-making. We have shown machine learning models can achieve validation PRC AUC scores ranging from 0.995 to 0.907 across targeted hematological and solid tumour assays. The ability to classify clinically reportable variants provides decision support to manual expert curation, a significant bottleneck in scaling up clinical sequencing operations. Machine learning models allow the analysis of large numbers of variant annotations that can be complementary, redundant and sometimes conflicting. It allows for a more thorough interpretation of a variant’s consequence within a patient’s clinical context, a task that is demanding of trained genetic scientists.

These models’ provision of explanatory waterfall plots for individual variant’s prediction further aids scientists in evaluating and understanding the contributing factors to each decision, resulting in confidence in the decision support algorithms and the reporting process. The integration within the tertiary analysis workflow provides the much needed transparency of AI into critical healthcare systems. The integration also allows for future work to measure clinical efficiencies of implementing ML models and collect valuable feedback of scientists working on variant analysis. Another area for future study is the generalisation of the models across multiple assays and between laboratories to broaden the benefits of machine learning. The application of AI into the clinical reporting workflows has just begun and will allow the streamlining the reporting process and reduce inter-reviewer variability to allow for more accurate, robust and clinically defensible oncology reporting.

## Data Availability

All data produced in the present study are available upon reasonable request to the authors

## SUPPLEMENTARY METHODS

For information regarding the diagnostic service please see https://www.petermac.org/molecular-pathology

### Haem Assay and Myeloid Assay – QIAseq custom design

Targeted sequencing of 57 genes was performed using custom QIAGEN QIAseq single primer extension-based target enrichment and Illumina NextSeq500 with 150 bp paired end read sequencing. A customised CLC Genomics Workbench (QIAGEN) analysis pipeline (v3.2) was used to generate aligned reads and call variants (single nucleotide variants and short insertions or deletions) against the hg19 human reference genome. Variants were analysed using PathOS software (Peter Mac) and described according to HGVS nomenclature. The ‘Haem’ assay included assessment of the full gene panel (n = 57 genes) and the ‘Myeloid’ panel included assessment of a limited set of either eight genes (between October 2021 and June 2022) or 22 genes (between July 2022 and December 2023).

### Solid Tumour Assay - TSO500

Targeted sequencing of 523 cancer genes from DNA and 55 cancer genes from RNA was performed using the Illumina TruSight™ Oncology 500 assay (TSO500). DNA and RNA were extracted from pathologist-selected areas of submitted formalin-fixed paraffin embedded (FFPE) tumour using the Qiagen AllPrep DNA/RNA FFPE kit. RNA was reverse transcribed to cDNA. DNA was ultra-sonically sheared to an appropriate size range. A ‘Unique Molecular Identifier’ (UMI) tagged DNA library and a standard RNA library were prepared and enriched with magnetic streptavidin beads following targeted hybridisation to gene-specific biotinylated probes. Pooled, normalised libraries were sequenced to an appropriate target mean coverage on an Illumina sequencing platform (NextSeq or NovaSeq). Illumina Software TSO500 v2.2 Local App was used to generate aligned reads and call variants against the hg37 human reference genome. Clinical Genomics Workspace (CGW) from PierianDx and Navify Mutation Profiler from Roche were used to annotate, filter and report clinically relevant findings.

## ABBREVIATIONS

API: Application Programming Interface
AIPA: AI prediction algorithm
AUC: Area Under the Curve
BAM: Binary Alignment Map format
CADD: Combined Annotation Dependent Depletion
cDNA: complementary DNA
CDSS: Clinical Decision Support System
delins: A variant which combines a deletion and an insertion
FDA: U.S. Food and Drug Administration
FN: False negatives
FP: False positives
HGVS: Human Genome Variant Society indel Insertion / Deletion
MNP: Multi-nucleotide polymorphism
MRD: Measurable Residual Disease
NGS: Next Generation Sequencing
NATA: National Association of Testing Authorities
NGS: Next Generation Sequencing
PCR: Polymerase Chain Reaction
PRC: Precision Recall Curve
ROC: Receiver Operating Characteristics
TP: True positives
TSV: Tab separated variable format
UMAP: Universal Manifold Approximation and Projection
UMI: Unique Molecular Identifier
VAF: Variant Allele Frequency
VCF: Variant Call Format

## DECLARATIONS

### Ethics approval and consent to participate

Ethics approval was granted on 1 April 2022 (EPIC Study Code: PMC81837, HREC: 81837).

### Consent for publication

Not applicable.

### Competing interests

The authors declare that they have no competing interests.

### Funding

This research was supported by the Laby Foundation, The Peter Mac Foundation, Therapeutics Innovation Australia and a National Health and Medical Research Council (NHMRC) Program Grant (1054618). The research benefitted by support from the Victorian State Government Operational Infrastructure Support and Australian Government NHMRC Independent Research Institute Infrastructure Support.

## Authors’ contributions

KDD conceived the study. KDD and RP wrote the software and wrote the manuscript. Ongoing feedback and advice given by AF, YK, RL, PB and SBF. All authors read and approved the final manuscript.

## Acknowledgements

The authors would like to acknowledge the generosity of our funders in making this project possible.

## Authors’ information (optional)

Not applicable.

## Notes

### Competing Interest Statement

The authors have declared no competing interest.

### Author Declarations

The Human Research Ethics and Governance Office (HREC) of the Peter MacCallum Cancer Centre granted Ethics approval on 1 April 2022 (EPIC Study Code: PMC81837, HREC: 81837).

